# Nomenclature for tracking of genetic variation of seasonal influenza viruses

**DOI:** 10.64898/2025.12.06.25341755

**Authors:** Richard A. Neher, John Huddleston, Trevor Bedford, Nicola S. Lewis, Ruth Harvey, Monica Galiano, Alexander M. P. Byrne, Sarah James, Derek Smith, Marta Łuksza, Denis Ruchnewitz, Michael Laessig, Seiichiro Fujisaki, Shinji Watanabe, Hideki Hasegawa, Norman Hassell, David E. Wentworth, Rebecca Kondor, Yi Mo Deng, Clyde Dapat, Kanta Subbarao, Ian Barr

**Affiliations:** Biozentrum, University of Basel, Basel, Switzerland; Swiss Institute of Bioinformatics, Basel, Switzerland; Vaccine and Infectious Disease Division, Fred Hutchinson Cancer Research Center, Seattle, WA, USA; Molecular and Cell Biology, University of Washington, Seattle, WA, USA; WHO Collaborating Centre for Reference and Research on Influenza, Crick Worldwide Influenza Centre, The Francis Crick Institute, London, UK; Center for Pathogen Evolution, Department of Zoology, University of Cambridge, Cambridge CB2 3EJ, UK; The Halvorsen Center for Computational Oncology, Department of Epidemiology and Biostatistics, Memorial Sloan Kettering Cancer Center, New York, NY, USA; Institute for Biological Physics, University of Cologne, Zülpicherstr. 77, 50937 Köln, Germany; Center for Predictive Analysis of Viral Evolution (PREVIR); Influenza Research Center, National Institute of Infectious Diseases, Japan Institute for Health Security, Tokyo, Japan; National Center for Immunization and Respiratory Diseases (NCIRD), Centers for Disease Control and Prevention (CDC), 1600 Clifton Road, Atlanta, GA 30333, USA; The WHO Collaborating Centre for Reference and Research on Influenza, The Peter Doherty Institute for Infection and Immunity, Melbourne, VIC, Australia; Department of Microbiology and Immunology, The University of Melbourne, The Peter Doherty Institute for Infection and Immunity, Melbourne, VIC, Australia

## Abstract

**Background:** Genomic surveillance of human seasonal influenza viruses is an essential component of the Global Influenza Surveillance and Response system (GISRS) and informs the recommendations for the seasonal influenza vaccine composition. Phylogenetic analysis of viral genome sequences is used to identify groups of viruses sharing potential antigenic change and computational models are used to predict which viral variants are likely to circulate at high levels in upcoming seasons. To facilitate discussion and reporting of genetic diversity, as well as to communicate antigen recommendations, up-to-date and sufficiently granular definitions of genetic clades are important.

**Methods:** A nomenclature system for segments 4 (haemagglutinin) and 6 (neuraminidase) of human A(H3N2), A(H1N1)pdm09, and influenza B.

**Results:** We devised a clade suggestion algorithm that proposes new subclades based on criteria including (i) the number of sequences in the group, (ii) the divergence from the directly ancestral clade, and (iii) the number and quality of amino acid substitutions on the branch leading to the common ancestor of the subclade. Algorithmic clade proposals were reviewed and assigned a systematic hierarchical label consisting of a leading letter, followed by numbers (e.g., G.1.3). Names are kept short by aliasing, that is collapsing prefixes into unique letters. Subclade definitions are shared openly to promote adoption and tool development. Nextclade is supporting this new nomenclature and it is being used routinely by the GISRS network.

**Conclusions:** With increasing genomic surveillance, the need for up-to-date classification schemes is growing and we hope that the current dynamic proposal will adapt to growing data volumes and aid in simplifying the interpretation of these data.

## I. INTRODUCTION

Seasonal human influenza viruses rapidly accumulate amino acid substitutions that change their antigenic properties, necessitating frequent updates of vaccines (Petrova and Russell, 2018). Recommendations for vaccine composition are made twice a year for the Southern and Northern Hemispheres, by a WHO-convened expert group, whose decisions are based on genetic and antigenic information generated at WHO influenza Collaborating Centers and National Influenza Centers (NIC’s) obtained from viruses collected through the GISRS network (Ziegler *et al*., 2018). Genomic surveillance of the four different seasonal influenza virus lineages A(H3N2), A(H1N1)pdm09, B/Victoria, B/Yamagata (not observed since April 2020) has become an important component of vaccine antigen composition recommendation. Genetic sequence data of influenza viruses, especially from the haemagglutinin (HA) gene segment and the neuraminidase (NA) gene segment, which code for the two key surface glycoproteins of the influenza A and B viruses that evolve rapidly because they are targeted by human immune response. The HA and NA glycoproteins are critical for entry and dissemination while antibodies that inhibit/neutralize these activities are important for protection from disease. Tracking frequencies of different viral HA and NA sequence variants allows quantification of geographic differences in circulation, and detection of substitutions with putative phenotypic consequence at much higher resolution than traditional serological assays. Furthermore, genetic sequences form the basis of more recent efforts to model and predict the frequencies of emerging variants within the viral population (Morris *et al*., 2018) including viruses showing reduced susceptibility to influenza antiviral drugs that target the NA or polymerase acidic (PA) proteins.

To effectively discuss the circulating diversity of HA and NA gene segments, characterize their geographic spread, and estimate their growth rates, it is essential to have a shared nomenclature to refer to and track relevant groups of genetically related viruses. Since individual gene segments of influenza viruses reassort during co-infections but don’t recombine efficiently, their evolutionary history is well described by phylogenetic analysis defining a hierarchical relationship between variants that naturally defines groups of similar viruses. GISRS uses such phylogenetic analysis to define clades, monophyletic groups of HA gene segments in the tree, that typically form groups of closely related viruses. These clades are named using alternating combinations of letters and numbers separated by periods (e.g. 3C.2a1b.2a.2). Similar nomenclature systems are also used for influenza viruses in animals, most prominently for avian H5 viruses (Shepard *et al*., 2014; Smith *et al*., 2015; World Health Organisation/World Organisation for Animal Health/Food and Agriculture Organisation H5N1 Evolution Working Group, 2008) and swine influenza viruses (Anderson *et al*., 2016).

In contrast to the animal influenza viruses, the existing nomenclature for human seasonal viruses has never been formalized and, in its current format, has a number of short-comings: As the viral population continues to evolve, clade names get longer and longer to track provenance within the name. Ad hoc shortening of names by stripping shared prefixes can result in ambiguities. At the same time, long names result in reluctance to designate new clades to avoid further lengthening names. But without continuously designating new names for emerging groups, the nomenclature quickly fails to break down circulating diversity in an informative and manageable way. Once most circulating viruses belong to the same clade, the nomenclature system is no longer useful for surveillance purposes.

Here, we use a dynamic nomenclature for naming genetic groups that will offer greater granularity than the previous clade nomenclature system while being unambiguous and easy to update. This system is similar to the PANGO nomenclature developed to track SARS-CoV-2 evolution, where the trade-off between long names and high granularity was solved by “aliasing”, that is the collapsing of long prefixes into unique abbreviations (Rambaut *et al*., 2020). The subclade nomenclature system was developed for the segments 4 and 6 of the influenza virus genome, coding for the HA and NA glycoproteins which are the most relevant for vaccine antigen selection. To ensure transparent communication of this nomenclature and to facilitate implementation of the scheme in software, we provide human and machine readable definitions on GitHub where they can be continuously updated and a record of all previous versions will be kept. Below, we separately discuss (i) how new subclades are defined and (ii) how they are named to form the basis of the new nomenclature system for influenza A and B virus HA and NA gene segments. These newly defined genetic groups will be called “subclades” to differentiate them from the existing clade nomenclature.

## II. METHODS

### A. Criteria of subclade definition

Rather than retrospectively breaking up a set of fixed sequences into genetic groups, we need a system that can designate new subclades as soon as new data becomes available. Once a particular genetic group meets certain criteria, it would be designated as a new clade with a unique name. We propose to designate new clades using a semi-automatic procedure that takes as input a phylogenetic tree annotated with existing clades, similar to tools developed for SARS-CoV-2 (McBroome *et al*., 2024). The phylogenetic trees maintained by Nextstrain (Hadfield *et al*., 2018) are used for this purpose, as these trees aim to be geographically representative, cover the last two years of global circulating influenza diversity, and are updated weekly. The exact tree and other parameters of the algorithm are specified in the file ‘config/suggestion-parameters.json’ in the same git repository documenting the subclade designations.

We defined a workflow that suggests a new clade designation based on three criteria:

- **Size and dynamics of the subclade:** big groups should have a higher priority for designation, especially if they recently expanded.
- **Divergence:** the more nucleotide substitutions have accumulated relative to the break point of the parent subclade, the higher the priority of designating a new subclade
- **Specific amino acid substitutions:** ideally, breakpoints sit on long branches with significant substitutions that might be of functional importance.

These criteria are quantified and combined into a single numerical score as detailed below.

#### Size and phylogenetic structure

To single out subclades in phylogeny that are of sufficient size and are phylogenetically distinct, we propose a measure like the Local Branching Index (LBI) (Neher *et al*., 2014) which tends to score polytomies (internal nodes on the phylogenetic tree with many direct children) highly. The score ϕ_n_ of node *n* is recursively calculated from the scores of its children *c* in one post-order traversal: 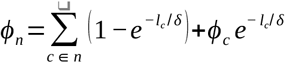 Here *l*_*c*_ is the length of the branch leading to child node *c* and *δ* is a distance scale determining how rapidly this score is “forgotten” along the tree. Terminal nodes have *ϕ*_*c*_ = 1. This way, the contribution of each terminal to its parent 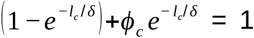 regardless of its branch length. Each internal branch has a maximum contribution of 1 and less than 1 if the branch is short compared to *δ*. In addition, we calculate the number of leaves below each branch in the tree, which is later used to ensure that every newly proposed subclade is larger than a minimal size. A region-resolved analysis of growth competition for subclades can serve to mitigate biases due to sampling inhomogeneities and seasonal growth variation (Meijers & Ruchnewitz *et al*., 2025)

#### Divergence

The measure of divergence used here is a count of nucleotide substitutions along the tree since the last subclade breakpoint. We selected nucleotide changes to more closely represent time, but amino acid substitutions could be used as well. The more divergence has accumulated since the last break point, the more readily a new subclade should be designated. The divergence score is called *d*_*n*_ below.

#### Branches with relevant substitutions

To prioritize branches with important substitutions and to pick branches with many nonsynonymous substitutions over those with only synonymous changes, each substitution is assigned a weight, and the sum of these weights is calculated for all substitutions on the branch. For A(H3N2) HA, this weight is currently 3 for each of the key sites identified by Koel *et al*., 2013, 2 for epitope sites (Bush *et al*., 1999), 1 for other HA1 or HA2 substitutions, and 0 for synonymous substitutions. For A(H1N1)pdm09, positions homologous to epitopes defined by Caton *et al*., 1982 are assigned weight 2, all other amino acid substitutions weight 1, and synonymous substitutions weight 0. For HA of influenza B and all neuraminidase segments, amino acid substitutions carry weight 1 and synonymous substitutions weight 0. These weights can be specified in a position specific way for each subtype or lineage for each segment. The branch score is called *b*_*n*_ below. The current version requires a branch score greater than zero, that is branches with only synonymous substitutions are not considered as breakpoints to define a new clade or subclade.

### B. Aggregation of scores

Above, three scores were defined that together should determine whether a branch should be designated the root of a new clade. The *branch score* and the *divergence score* essentially count substitutions and increase with the depth of the tree, but are fairly independent of sampling density. However, the phylogenetic bushiness *ϕ*_*n*_ and the size of clades depend strongly on sampling density. To consistently combine these into a single score, we need to normalize them. The suggested normalization for the divergence, the branch score, and the bushiness score is linear saturation. If *x* is the raw score, the normalized score *X is calculated* as

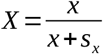

where *s*_*x*_ is a scale associated with the score. When *x*=*s*_*x*_, the score equals 0.5 and it saturates at 1 for large values of *x*. For the branch score and the divergence, the scale is essentially given by the number of substitutions considered as significant. Since the phylogenetic score depends on the sample size, it makes sense to choose that normalization be proportional to the size of the tree. We define *s*_*ϕ*_ to be the median value of *ϕ* across the tree.

### C. Clade designations

New clades are assigned by walking through the tree in pre-order (parents before children) and a new clade is designated if the sum of the normalized scores exceeds a certain threshold *θ*.

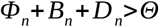

The current value for the threshold in the influenza virus lineage assignment is *θ* = 1.0. This means any one score alone is insufficient to trigger a new lineage since all normalized scores are below 1.0 by definition. But together with a phylogenetic score and the divergence contribution, the threshold can be crossed for a single HA1 mutation. Note that the divergence *D*_*n*_ of the downstream part of the tree needs to be recalculated after designating a new subclade since divergence is relative to the parent clade. In addition to the threshold condition defined above, we require the subclades to be larger than a minimal size and the branch to have at least one amino acid substitution. This minimal size is currently set at 30 genomes in the 2y trees from Nextstrain. Subclades that meet these criteria are considered “proposals” for manual review by WHO Collaborating Centres for Influenza and academic groups supporting the vaccine antigen selection process. In addition, clades of particular interest can be added manually outside of the suggestion algorithm.

This algorithm is implemented in python and the code is available in the GitHub repository <u>influenza-clade nomenclature/clade-suggestion-algorithm</u>. The threshold parameters for the different subtypes, lineages, and segments are specified in configuration files in the associated GitHub repositories for the individual nomenclatures and differ slightly between segments.

### D. Subclade Naming and Implementation for Seasonal Influenza Viruses

The system uses a variation of the Pango nomenclature system developed for SARS-CoV-2 (Rambaut *et al*., 2020). As in the Pango scheme, we use capital letters as aliases and numbers separated by periods for the retained hierarchical part of the name. Shortening or aliasing of the name typically happens after three hierarchical levels, but in contrast to Pango we do not mandate aliasing after a fixed length. Sometimes, it might be appropriate to alias earlierif there is a rapid lineage expansion, lineage replacement, or significant genetic changes likely to affect virus phenotype. Subject matter experts can propose to override the default naming to simplify communications and highlight important emerging lineages. After the introduction of an alias (e.g., ‘D’ for ‘C.1.1.1’), both ‘D’ and ‘C.1.1.1’ are considered valid names for the group. For aliasing purposes, the letter H and N will be excluded to avoid confusion with the influenza A subtype nomenclature, while I and O will be avoided since they are often hard to distinguish from numbers one and zero.

## III. RESULTS

This nomenclature system was implemented for the HA and NA segments of A(H3N2), A(H1N1)pdm09, and B/Vic. For HA, we initialized the nomenclature by mirroring existing nomenclature conventions starting at clade 3C for A(H3N2) (around 2012), 6.1A.5a for A(H1N1)pdm09 (around 2018), and V1A for B/Vic (around 2008). The traditional clade nomenclature for seasonal influenza viruses has not been updated since early 2023, but we have kept adding new subclades using the suggestion algorithm described above followed by manual review by us. The subclades defined at the time of writing for the HA segments of the three currently circulating seasonal lineages are shown in Fig. 1 and summarized in supplementary tables 1-3. Aliases A-D are used for A(H1N1)pdm09, A-K for A(H3N2), and A-C for B/Vic. No analysis was performed for B/Yam viruses as there are no recorded HA sequences available since March 2020 and it appears that this B lineage is no longer circulating in any part of the world (Caini *et al*., 2024).

**FIG. 1.**
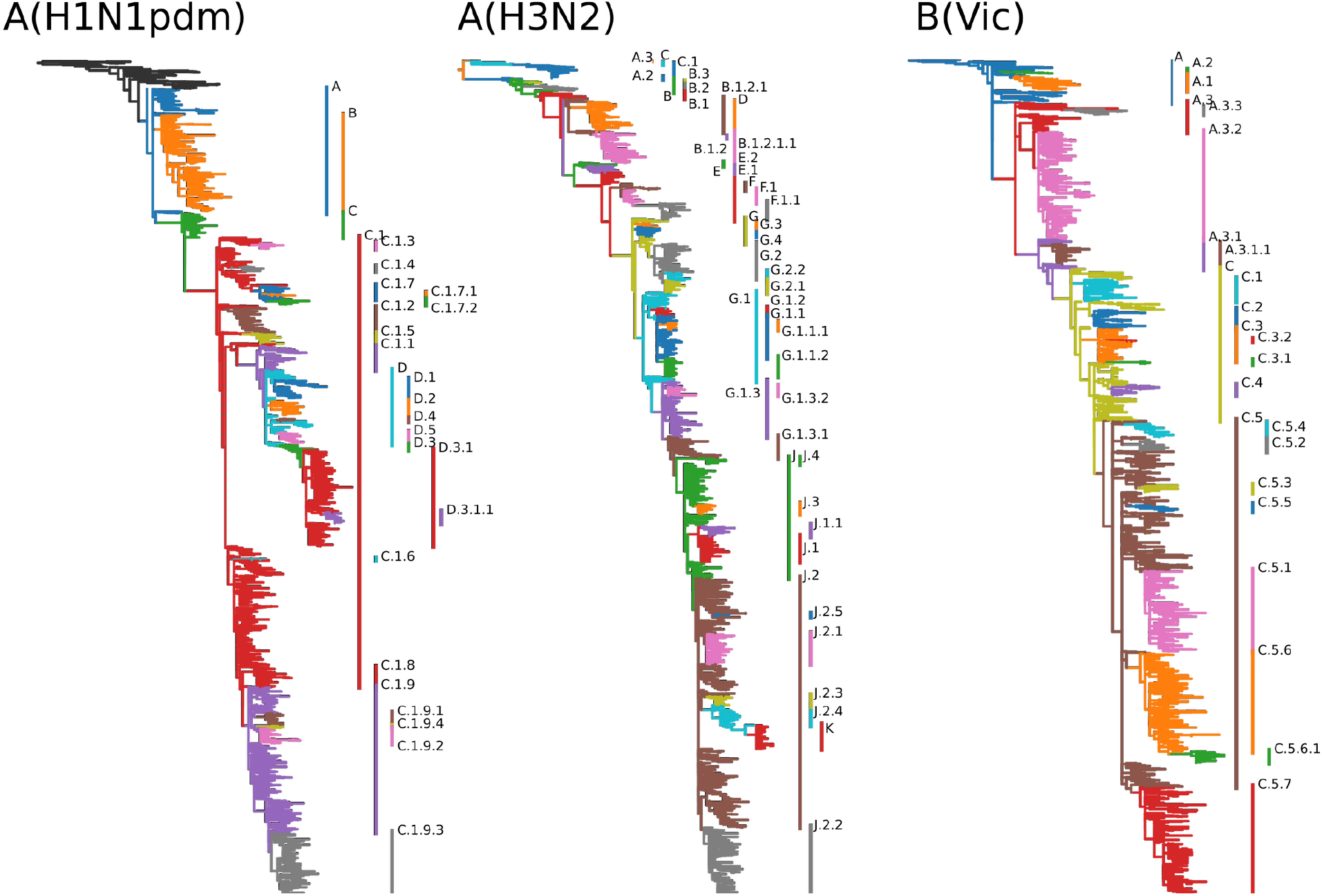
Proposed subclade nomenclature for HA segments of currently circulating seasonal human influenza viruses as annotated on Nextstrain’s continuously updated analysis.

For NA, the nomenclature was initialized at a prominent branch ancestral to currently circulating diversity and subclades were added after manual review from suggestions by the algorithm as described above. The resulting clades are shown in Fig. 2 and summarized in supplementary tables 4-6. Frequent convergent evolution of NA can lead to unstable trees and occasionally results in sister clades nested in each other, as is the case for the NA clade B.3 of A(H3N2).

**FIG. 2.**
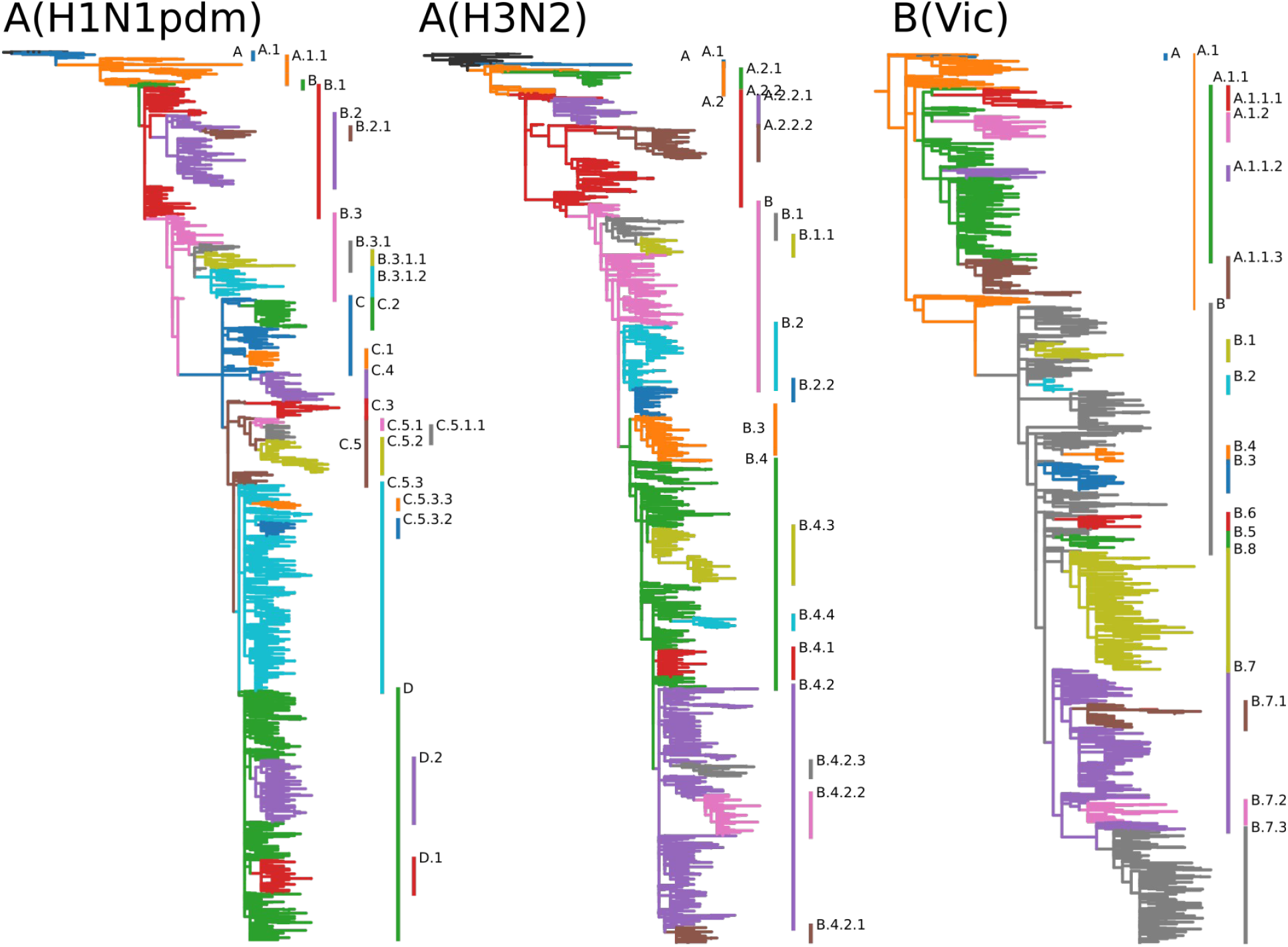
Proposed subclade nomenclature for NA segments of currently circulating seasonal human influenza viruses as annotated on Nextstrain’s continuously updated analysis.

For each of the three current lineages, subclades break up circulating diversity and allow easy tracking of relative frequencies of variants in different parts of the world. The lineage definitions are available in git repositories – one for each lineage and segment – in the GitHub organization <u>influenza-clade-nomenclature</u>. Each clade or subclade is defined via a machine-readable configuration file that contains the name, parent, and defining substitutions of the clade. Optionally, the definition can contain a list of representative sequences. These individual definitions are automatically converted into human readable files summarizing all subclades. Currently defined subclades for the HA and NA segements of the lineages A(H1N1pdm), A(H3N2), and B(Vic) are provided as supplementary tables S1-6.

The current plan is to review and potentially update the nomenclature four times a year at the beginning and the end of the influenza seasons in the Northern and Southern Hemispheres. Updates are logged in the Git repositories at github.com/influenza-clade-nomenclature and accompanied by a short summary of the update.

The aliasing procedure is an essential component of the nomenclature system to keep clade names short and to facilitate discussions of viral diversity. Fig. 3 illustrates this aliasing procedure using a recent example of A(H3N2) HA evolution. The currently dominating A(H3N2) HA subclade K is an alias for J.2.4.1, whose ancestors is variant J.2.4.

**Fig. 3.**
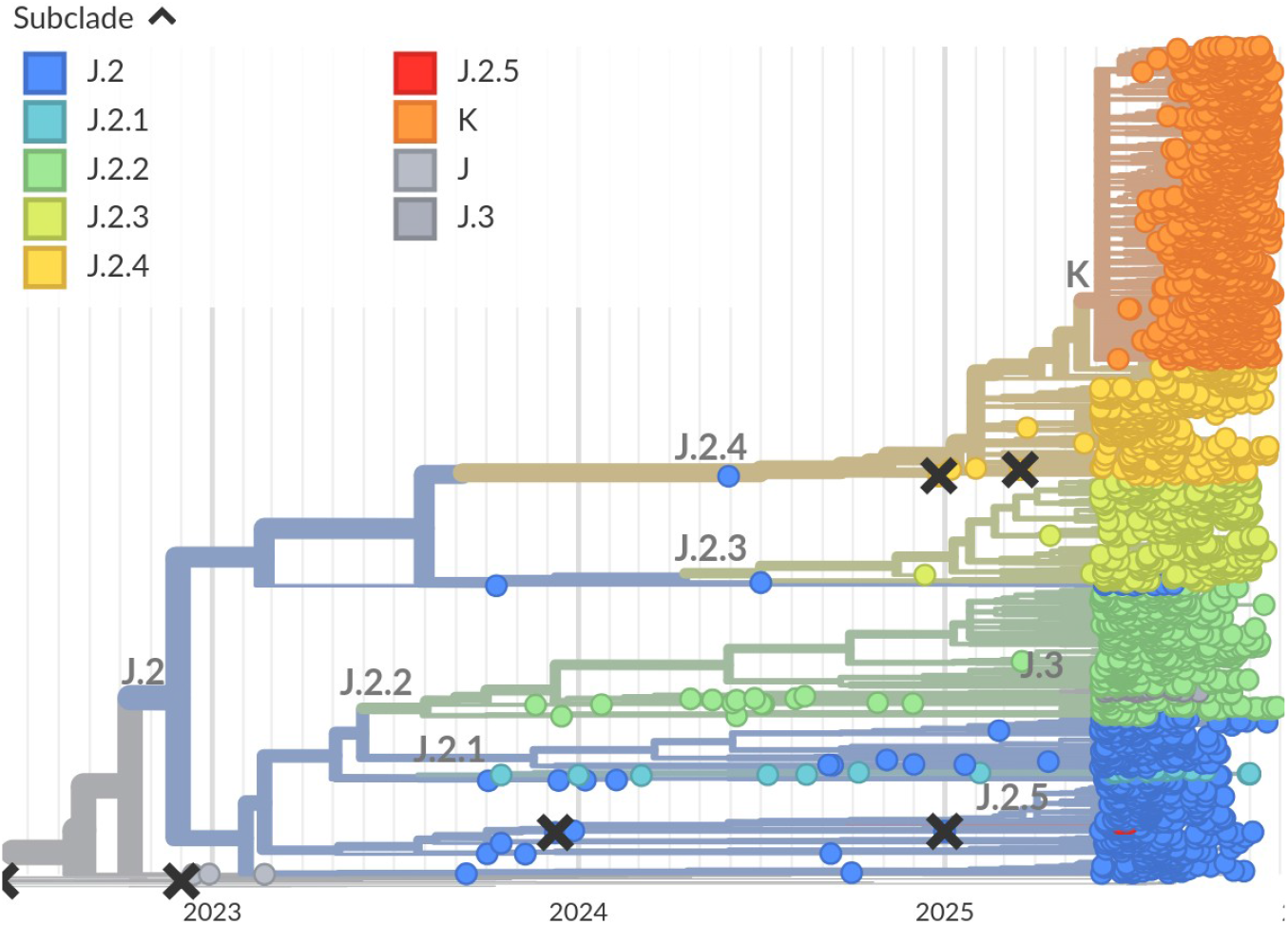
A group of viruses in subclade J.2.4 of HA A(H3N2), highlighted in orange, acquired several substitutions and rapidly rose in frequency in the second half of 2025. This group was designated J.2.4.1 and an alias K was introduced to facilitate discussion of this subclade, which will likely dominate the 2025/26 Northern Hemisphere season (Figure from nextstrain.org).

The nomenclature scheme is implemented in Nextclade datasets maintained by the Nextstrain team (Aksamentov *et al*., 2021). Nextclade has a web interface that allows users to rapidly assign clades and subclades to their sequences. To assign clades to sequences, a user can simply drop a file with HA or NA sequences from one subtype or lineage on the interface, select a reference dataset, and run the analyses. Analyzing 100 sequences takes a few seconds, input data not matching the reference data set will be ignored. A command line interface (CLI) is available for the analysis of large quantities of data and database updates (it takes about one minute to classify 100,000 HA sequences on a laptop).

## IV. CONCLUSION

The unprecedented genomic surveillance efforts for SARS-CoV-2 (currently >17 million sequences on GISAID (Shu and McCauley, 2017)) have highlighted the importance of dynamic nomenclature schemes to track viral variants (Rambaut *et al*., 2020). Along with SARS-CoV-2, sequencing volumes for other viruses is also increasing steeply, including seasonal influenza viruses with >400k HA and NA sequences in GISAID. Systematic nomenclature systems have recently been proposed for RSV (Goya *et al*., 2024) and MPXV (Happi *et al*., 2022). Similar to these proposals, we propose a nomenclature scheme that shortens hierarchical names by collapsing prefixes into aliases. We also adopted GitHub as a place to disseminate the nomenclature and document updates to the nomenclature. Our proposal differs from the above in that aliasing is not automatic, but can be triggered after 2, 3, or 4 hierarchical levels depending on the anticipated utility of the alias. The rapid evolution of HA of A(H3N2) viruses provides a recent example where aliasing proved useful (see Fig. 3): Subclade J had dominated since mid 2023 and was split into J.1-4 in late 2023. From late 2024 onward, J.2 was split further into J.2.1-5. The observation of a variant in J.2.4 with several significant mutations triggered the designation of J.2.4.1 in Oct 2025, for which an alias K was introduced two weeks later. This alias significantly facilitates discussion of this group of viruses, which will likely require further subdivision into subclades K.x soon. Furthermore, we propose a hybrid clade designation process where an algorithm objectively proposes new clades (similar to autolin, (McBroome *et al*., 2024)), and experts review these proposals to ensure the nomenclature meets the needs of the GISRS community.

The system uses common rules for designating and naming genetic groups as subclades across all seasonal influenza virus lineages for their segments 4 (HA) and 6 (NA). At present, these subclades are mostly used to track emerging variants of the HA segment, but nomenclature for NA is increasingly used to describe reassorted genomic constellations (see Figure S1 for a recent example in A(H1N1)pdm09). Going forward, this system could be extended to additional gene segments though segments coding for internal proteins of the virus are more conserved and of lower relevance for the vaccine selection process. Given the similarity of the clade names across lineages and segments, the clade names are prefixed with the segment and subtype (e.g. H3-J.2.5) if the relevant lineage and segment are not clear from context.

Classification schemes for viral diversity are most useful when they are widely used by the community, and if they are kept up-to-date and continue to delineate relevant groups of circulating diversity. We hope that with this dynamic influenza A and B HA and NA subclade nomenclature will meet the scientific community’s needs, and this note is primarily about the process of updating and disseminating the nomenclature rather than the clade definitions themselves. This nomenclature system has been utilized by the GISRS system for the last 2 years and has proved very useful. It has also been embraced by other parties such as influenza vaccine manufacturers and researchers. The infrastructure to maintain, update, and disseminate the scheme is designed to be open, dynamic, and agnostic of downstream use. The community is encouraged to build tools around it and contribute to the maintenance of the nomenclature and to use this nomenclature in reports and publications.

## Data Availability

No new data was generated as part of this study. The dynamic nomenclature described here is available at
https://github.com/orgs/influenza-clade-nomenclature

## Acknowledgements

We gratefully acknowledge stimulating discussions with members of the GISRS community. This work was supported by core funding from the Francis Crick Institute from Cancer Research UK, the UK Medical Research Council, and the Wellcome Trust.

