## Supplementary text and tables for "Nomenclature for tracking of genetic variation of seasonal influenza viruses"

#### Supplementary Materials

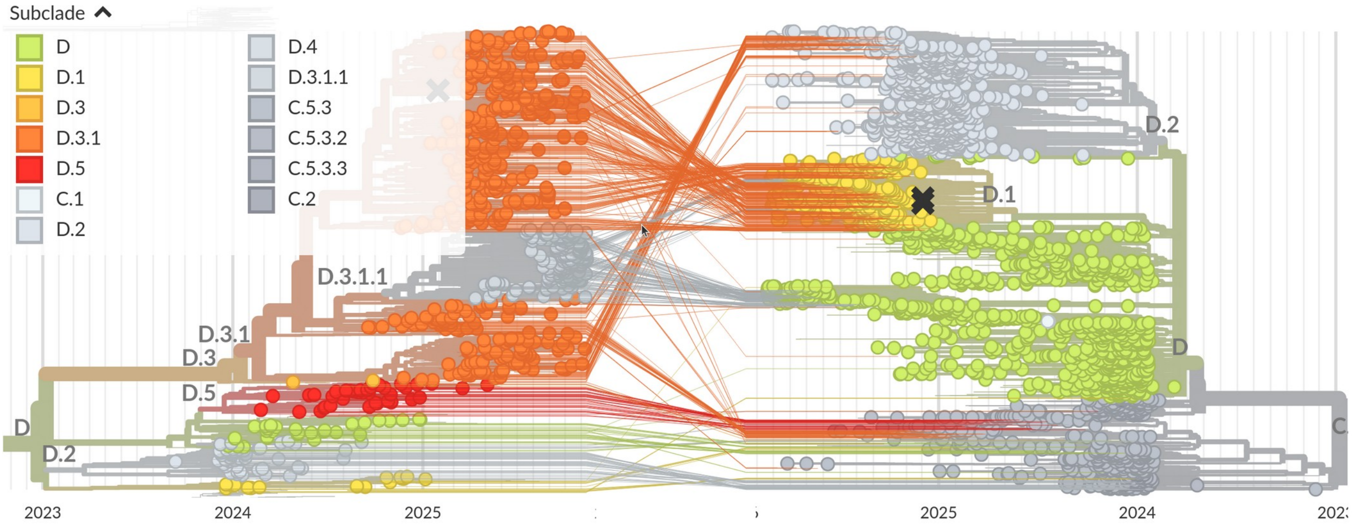


Supplementary figure 1: Tangle-gram of recent H1N1pdm isolates focusing on subclade H1-D of HA on the left with the Neuraminidase tree on the right. Subclade nomenclatures for both segments allows for straightforward identification of mulit-segment constellations.

### Tables

TABLE S1 A(H3N2) HA clade definitions

| Subclade | Clade | full subclade name |
|---|---|---|
| A | 3C | A |
| A.2 | 3C.2 | A.2 |
| A.3 | 3C.3 | A.3 |
| A.3.2 | 3C.3b | A.3.2 |
| B | 3C.2a | A.2.1 |
| B.1 | 3C.2a1 | A.2.1.1 |
| B.1.1 | 3C.2a1a | A.2.1.1.1 |
| B.1.2 | 3C.2a1b | A.2.1.1.2 |
| B.1.2.1 | 3C.2a1b.1 | A.2.1.1.2.1 |
| B.1.2.1.1 | 3C.2a1b.1a | A.2.1.1.2.1.1 |
| B.2 | 3C.2a2 | A.2.1.2 |
| B.3 | 3C.2a3 | A.2.1.3 |
| B.4 | 3C.2a4 | A.2.1.4 |
| C | 3C.3a | A.3.1 |
| C.1 | 3C.3a1 | A.3.1.1 |
| D | 3C.2a1b.1b | A.2.1.1.2.1.2 |
| E | 3C.2a1b.2 | A.2.1.1.2.2 |
| E.1 | 3C.2a1b.2a | A.2.1.1.2.2.1 |
| E.2 | 3C.2a1b.2b | A.2.1.1.2.2.2 |
| F | 1 | A.2.1.1.2.2.1.1 |
| F.1 | 1a | A.2.1.1.2.2.1.1.1 |
| F.1.1 | 1a.1 | A.2.1.1.2.2.1.1.1.1 |
| G | 2 | A.2.1.1.2.2.1.2 |
| G.1 | 2a | A.2.1.1.2.2.1.2.1 |
| G.1.1 | 2a.1 | A.2.1.1.2.2.1.2.1.1 |
| G.1.1.1 | 2a.1a | A.2.1.1.2.2.1.2.1.1.1 |

| | | |
|---|---|---|
| G.1.1.2 | 2a.1b | A.2.1.1.2.2.1.2.1.1.2 |
| G.1.2 | 2a.2 | A.2.1.1.2.2.1.2.1.2 |
| G.1.3 | 2a.3 | A.2.1.1.2.2.1.2.1.3 |
| G.1.3.1 | 2a.3a | A.2.1.1.2.2.1.2.1.3.1 |
| G.1.3.2 | 2a.3b | A.2.1.1.2.2.1.2.1.3.2 |
| G.2 | 2b | A.2.1.1.2.2.1.2.2 |
| G.2.1 | none | A.2.1.1.2.2.1.2.2.1 |
| G.2.2 | none | A.2.1.1.2.2.1.2.2.2 |
| G.3 | 2c | A.2.1.1.2.2.1.2.3 |
| G.4 | 2d | A.2.1.1.2.2.1.2.4 |
| J | 2a.3a.1 | A.2.1.1.2.2.1.2.1.3.1.1 |
| J.1 | none | A.2.1.1.2.2.1.2.1.3.1.1.1 |
| J.1.1 | none | A.2.1.1.2.2.1.2.1.3.1.1.1.1 |
| J.2 | none | A.2.1.1.2.2.1.2.1.3.1.1.2 |
| J.2.1 | none | A.2.1.1.2.2.1.2.1.3.1.1.2.1 |
| J.2.2 | none | A.2.1.1.2.2.1.2.1.3.1.1.2.2 |
| J.2.3 | none | A.2.1.1.2.2.1.2.1.3.1.1.2.3 |
| J.2.4 | none | A.2.1.1.2.2.1.2.1.3.1.1.2.4 |
| K | none | A.2.1.1.2.2.1.2.1.3.1.1.2.4.1 |
| J.2.5 | none | A.2.1.1.2.2.1.2.1.3.1.1.2.5 |
| J.3 | none | A.2.1.1.2.2.1.2.1.3.1.1.3 |
| J.4 | none | A.2.1.1.2.2.1.2.1.3.1.1.4 |

TABLE S2 A(H1N1)pdm09 HA clade definitions

| Subclade | Clade | full subclade name |
|---|---|---|
| A | 6B.1A.5a | A |
| B | 5a.1 | A.1 |
| C | 5a.2 | A.2 |
| C.1 | 5a.2a | A.2.1 |
| C.1.1 | 5a.2a.1 | A.2.1.1 |
| C.1.2 | none | A.2.1.2 |
| C.1.3 | none | A.2.1.3 |
| C.1.4 | none | A.2.1.4 |
| C.1.5 | none | A.2.1.5 |
| C.1.6 | none | A.2.1.6 |
| C.1.7 | none | A.2.1.7 |
| C.1.7.1 | none | A.2.1.7.1 |
| C.1.7.2 | none | A.2.1.7.2 |
| C.1.8 | none | A.2.1.8 |
| C.1.9 | none | A.2.1.9 |
| C.1.9.1 | none | A.2.1.9.1 |
| C.1.9.2 | none | A.2.1.9.2 |
| C.1.9.3 | none | A.2.1.9.3 |
| C.1.9.4 | none | A.2.1.9.4 |
| D | none | A.2.1.1.1 |
| D.1 | none | A.2.1.1.1.1 |
| D.2 | none | A.2.1.1.1.2 |
| D.3 | none | A.2.1.1.1.3 |
| D.3.1 | none | A.2.1.1.1.3.1 |
| D.3.1.1 | none | A.2.1.1.1.3.1.1 |
| D.4 | none | A.2.1.1.1.4 |
| D.5 | none | A.2.1.1.1.5 |

TABLE S3 B/Vic HA clade definitions

| Subclade | Clade | full subclade name |
|---|---|---|
| A | V1A | A |
| A.1 | V1A.1 | A.1 |
| A.2 | V1A.2 | A.2 |
| A.3 | V1A.3 | A.3 |
| A.3.1 | V1A.3a | A.3.1 |
| A.3.1.1 | V1A.3a.1 | A.3.1.1 |
| A.3.2 | none | A.3.2 |
| A.3.3 | none | A.3.3 |
| B | V1B | B |
| C | V1A.3a.2 | A.3.1.2 |
| C.1 | none | A.3.1.2.1 |
| C.2 | none | A.3.1.2.2 |
| C.3 | none | A.3.1.2.3 |
| C.3.1 | none | A.3.1.2.3.1 |
| C.3.2 | none | A.3.1.2.3.2 |
| C.4 | none | A.3.1.2.4 |
| C.5 | none | A.3.1.2.5 |
| C.5.1 | none | A.3.1.2.5.1 |
| C.5.2 | none | A.3.1.2.5.2 |
| C.5.3 | none | A.3.1.2.5.3 |
| C.5.4 | none | A.3.1.2.5.4 |
| C.5.5 | none | A.3.1.2.5.5 |
| C.5.6 | none | A.3.1.2.5.6 |
| C.5.6.1 | none | A.3.1.2.5.6.1 |
| C.5.7 | none | A.3.1.2.5.7 |

TABLE S4 A(H1N1)pdm09 NA clade definitions

| Subclade | full subclade name |
|---|---|
| A | A |
| A.1 | A.1 |
| A.1.1 | A.1.1 |
| B | A.1.1.1 |
| B.1 | A.1.1.1.1 |
| B.2 | A.1.1.1.2 |
| B.2.1 | A.1.1.1.2.1 |
| B.3 | A.1.1.1.3 |
| B.3.1 | A.1.1.1.3.1 |
| B.3.1.1 | A.1.1.1.3.1.1 |
| B.3.1.2 | A.1.1.1.3.1.2 |
| C | A.1.1.1.3.2 |
| C.1 | A.1.1.1.3.2.1 |
| C.2 | A.1.1.1.3.2.2 |
| C.3 | A.1.1.1.3.2.3 |
| C.4 | A.1.1.1.3.2.4 |
| C.5 | A.1.1.1.3.2.5 |
| C.5.1 | A.1.1.1.3.2.5.1 |
| C.5.1.1 | A.1.1.1.3.2.5.1.1 |
| C.5.2 | A.1.1.1.3.2.5.2 |
| C.5.3 | A.1.1.1.3.2.5.3 |
| C.5.3.2 | A.1.1.1.3.2.5.3.2 |
| C.5.3.3 | A.1.1.1.3.2.5.3.3 |
| D | A.1.1.1.3.2.5.3.1 |
| D.1 | A.1.1.1.3.2.5.3.1.1 |
| D.2 | A.1.1.1.3.2.5.3.1.2 |

TABLE S5 A(H3N2) NA clade definitions

| Subclade | full subclade name |
|---|---|
| A | A |
| A.1 | A.1 |
| A.2 | A.2 |
| A.2.1 | A.2.1 |
| A.2.2 | A.2.2 |
| A.2.2.1 | A.2.2.1 |
| A.2.2.2 | A.2.2.2 |
| B | A.2.2.3 |
| B.1 | A.2.2.3.1 |
| B.1.1 | A.2.2.3.1.1 |
| B.2 | A.2.2.3.2 |
| B.2.2 | A.2.2.3.2.2 |
| B.3 | A.2.2.3.3 |
| B.4 | A.2.2.3.4 |
| B.4.1 | A.2.2.3.4.1 |
| B.4.2 | A.2.2.3.4.2 |
| B.4.2.1 | A.2.2.3.4.2.1 |
| B.4.2.2 | A.2.2.3.4.2.2 |
| B.4.2.3 | A.2.2.3.4.2.3 |
| B.4.3 | A.2.2.3.4.3 |
| B.4.4 | A.2.2.3.4.4 |

TABLE S6 B/Vic NA clade definitions

| Subclade | full subclade name |
|---|---|
| A | A |
| A.1 | A.1 |
| A.1.1 | A.1.1 |
| A.1.1.1 | A.1.1.1 |
| A.1.1.2 | A.1.1.2 |
| A.1.1.3 | A.1.1.3 |
| A.1.1.4 | A.1.1.4 |
| B | A.1.2 |
| B.1 | A.1.2.1 |
| B.2 | A.1.2.2 |
| B.3 | A.1.2.3 |
| B.4 | A.1.2.4 |
| B.5 | A.1.2.5 |
| B.6 | A.1.2.6 |
| B.7 | A.1.2.7 |
| B.7.1 | A.1.2.7.1 |
| B.7.2 | A.1.2.7.2 |
| B.7.3 | A.1.2.7.3 |
| B.8 | A.1.2.8 |
